# Detection and Characterization of Human Bocavirus in Stool Samples from an Acute Gastroenteritis Outbreak in Salvador, Bahia, Brazil, in 2024

**DOI:** 10.64898/2026.10.19.26365916

**Authors:** Beatriz Peres Vidal, Silvana Oliveira Neves, Ively Paixão Santos, Silvia Ines Sardi, Gubio Soares Campos

## Abstract

**Introduction:** Acute gastroenteritis (AGE) is a global public health problem and a major cause of mortality in low- and lower-middle-income countries. Human bocavirus (HBoV) is among the viral agents associated with this condition. Therefore, investigating the presence of HBoV in cases of AGE in Salvador, Bahia, as well as its prevalence and circulating strains, is important.

**Objective:** To investigate the occurrence of HBoV infection during an AGE outbreak in Salvador.

**Methods:** A total of 178 samples were analyzed for HBoV using nested polymerase chain reaction (nested PCR), and the amplification products were visualized by agarose gel electrophoresis. HBoV was detected in 5.7% of the samples analyzed.

**Conclusion:** This study contributes to the understanding of HBoV epidemiology as a secondary agent associated with acute gastroenteritis during a norovirus outbreak in Salvador. Continued investigation and identification of the etiological agents involved in outbreaks are essential for evaluating the causes of AGE. These findings also highlight the need for broader diagnostic approaches and the importance of phylogenetic analysis for monitoring and controlling viral infections.

## Introduction

Acute gastroenteritis can require hospitalization in severe cases, as reported in previous studies (Cheng et al., 2008). One of the viral agents associated with this condition is human bocavirus (HBoV), first identified by Allander in Sweden in 2005 (Allander, 2008). HBoV is a single-stranded DNA virus belonging to the family Parvoviridae, subfamily Parvovirinae, and genus Bocaparvovirus, which infects several mammalian species, including humans (Lin et al., 2020). Among the HBoV types, HBoV-2, HBoV-3, and HBoV-4 have been associated with gastrointestinal disease (Allander, 2008; Lin et al., 2020).

HBoV has been reported in association with both acute gastroenteritis and respiratory tract infections. Given its respiratory tropism, HBoV infection has been associated with a greater risk of severe respiratory illnesses, including pneumonia and bronchiolitis (Mohammadi, 2023). Over time, HBoV has also been linked to other conditions, such as acute conjunctivitis (Johnson et al., 2023), myocarditis (Thanasia et al., 2023), and chronic otitis media (Runge et al., 2023).

In addition to tissue tropism, the association of HBoV with both gastrointestinal and respiratory infections may be related to its ability to undergo mutations despite being a DNA virus. It has therefore been proposed that an enteric HBoV lineage may have given rise to HBoV-1, which infects the respiratory tract (Lekana-Douki et al., 2018).

The HBoV genome consists of approximately 5,300 nucleotides of linear, single-stranded DNA with either positive or negative polarity. The virus encodes the structural proteins VP1, VP2, and VP3 (Li et al., 2015; Luo et al., 2021), which form its icosahedral capsid, and the nonstructural proteins NP1, NS1, NS1-70, NS2, NS3, and NS4 (Shen et al., 2015). HBoV is a nonenveloped virus (Guido et al., 2016). Although its replication cycle has not been fully characterized, some aspects of HBoV replication and replication mechanisms within the Parvoviridae family have been described (Knipe; Howley, 2013).

Human bocavirus infection triggers a complex immune response involving both T-cell-mediated immunity and humoral immunity through antibody production by B cells (Karaaslan et al., 2024). These responses are important in host defense against viral infections. HBoV capsid proteins, particularly VP1 and VP2, are targets of adaptive immune responses (Bhat; Almajhdi, 2021).

Previous studies have reported a high seroprevalence of IgG antibodies specific to the VP2 protein in the population (Karalar et al., 2010). Capsid-specific T-cell responses have also been detected, particularly in children. T cells play an important role in the immune response to HBoV. Helper T cells (CD4+) are prominent and contribute to the activation of cytotoxic T cells (CD8+) and natural killer (NK) cells, which participate in the elimination of infected cells (Karaaslan et al., 2024). Studies have highlighted the importance of CD4+ T-cell activation in antiviral immunity. In adults exposed to the virus, significant production of interferon gamma (IFN-γ), a cytokine involved in the Th1 response, has been observed, indicating a strong immune response.

Overall, the immune response to HBoV involves complex interactions between T and B lymphocytes, with T cells contributing substantially to viral clearance. Nevertheless, HBoV has mechanisms that may enable it to evade host immunity and facilitate replication. Further studies are needed to clarify these immune responses and the mechanisms underlying viral immune evasion (Bhat; Almajhdi, 2021).

Several studies have described HBoV as a causative agent of acute gastroenteritis (Campos et al., 2016; Lekana-Douki et al., 2018; Santos et al., 2010), with reported prevalence values reaching 53.8% (Bergallo et al., 2018; Cheng et al., 2008; Mohammad; Madi; Al-Nakib, 2020; Nora-Krukle et al., 2018).

## Materials and Methods

This study was approved by the Research Ethics Committee of the Institute of Health Sciences, Federal University of Bahia (UFBA), under CAAE registration number 61239422.9.0000.5662. This was a descriptive cross-sectional study involving 178 samples collected during an AGE outbreak in Salvador, Bahia, between May and September 2024. The samples were sent to the Virology Laboratory of the UFBA Institute of Health Sciences, where they were stored at ™80 °C. Stool supernatants were then prepared for viral nucleic acid extraction.

Nucleic acids were extracted from 140 µL of each stool supernatant using the QIAamp Viral DNA Kit® (QIAGEN, Hilden, Germany), following the manufacturer’s instructions. Viral DNA was analyzed by nested PCR using previously published methods and primers (Kapoor et al., 2011) and the GoTaq® G2 Colorless Master Mix (Promega, USA). An HBoV-2 strain (GenBank accession KM366092), previously detected in Brazil, was used as a positive control (Campos et al., 2016). Nested-PCR products were resolved by electrophoresis on 2% agarose gels prepared in TBE (Tris-borate-EDTA) buffer. The gels were stained with ethidium bromide (10 µg/mL) for 5 min and visualized under ultraviolet light. The expected HBoV amplification product was 576 bp, as previously reported (Kapoor et al., 2011).

Data were analyzed using Epi Info software, version 7.6.2.0. Fisher’s exact test was used to compare categorical variables. Statistical significance was set at p < 0.05.

## Results

Nested PCR was performed on 178 samples collected during a gastroenteritis outbreak at a private hospital in Salvador, Bahia. Of these, 10 samples (5.7%) tested positive for HBoV and 168 (94.3%) tested negative. The samples collected during the outbreak were assessed in relation to the reported AGE cases. The age distribution of patients with HBoV-positive AGE was also evaluated, and the authors reported that young adults were the most affected group.

Analysis of the nested-PCR products by agarose gel electrophoresis revealed a 576-bp amplicon (Figure 1). Three of the ten HBoV-positive samples are shown in the figure. Samples testing positive for HBoV by nested PCR were subsequently subjected to in vitro isolation and examination in Vero cell cultures. Cell culture enabled the assessment of cytopathic effects associated with HBoV infection in the host cells. The main cytopathic changes observed were cell rounding, clustering, and lysis (Figure 2). Immunofluorescence analysis was also performed on HBoV-infected cells (Figure 3). Viral titration was additionally performed, yielding a reported titer of 1 × 10^6.5.

**Figure 1.**
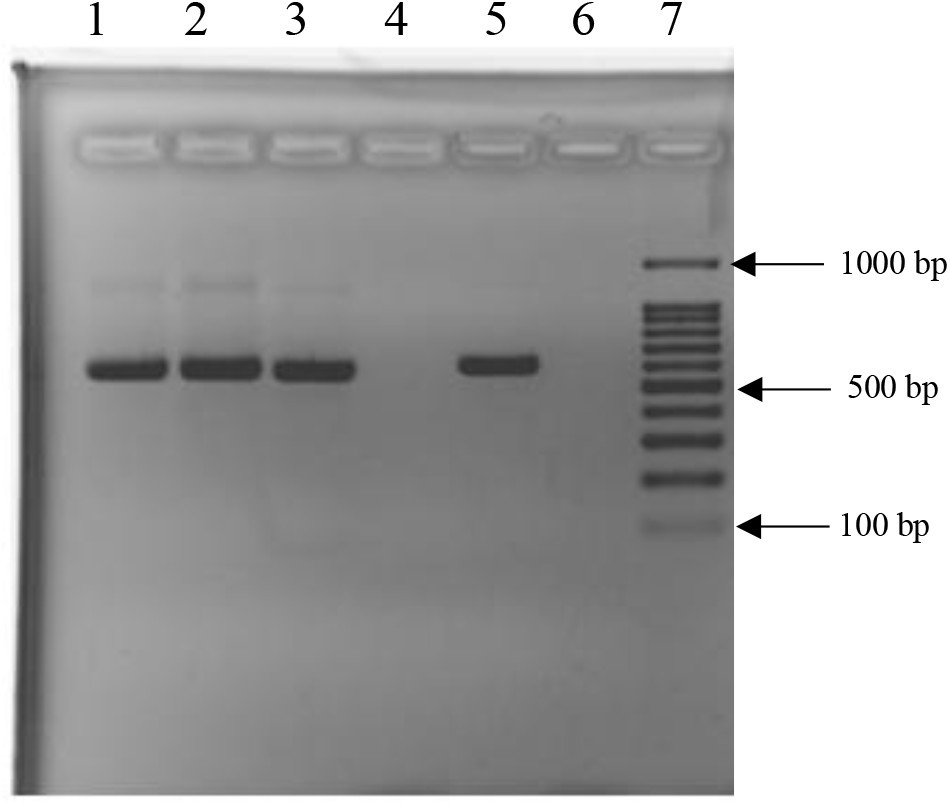
Detection of HBoV by nested PCR. A 2% agarose gel stained with ethidium bromide showing HBoV nested-PCR products. Lanes 1, 2, and 3: HBoV-positive samples. Lane 5: HBoV-positive control. Lanes 4 and 6: HBoV-negative samples. Lane 7: 100-bp DNA ladder (Invitrogen, USA). *Source: 1 Authors’ own work, 2025*.

**Figure 2.**
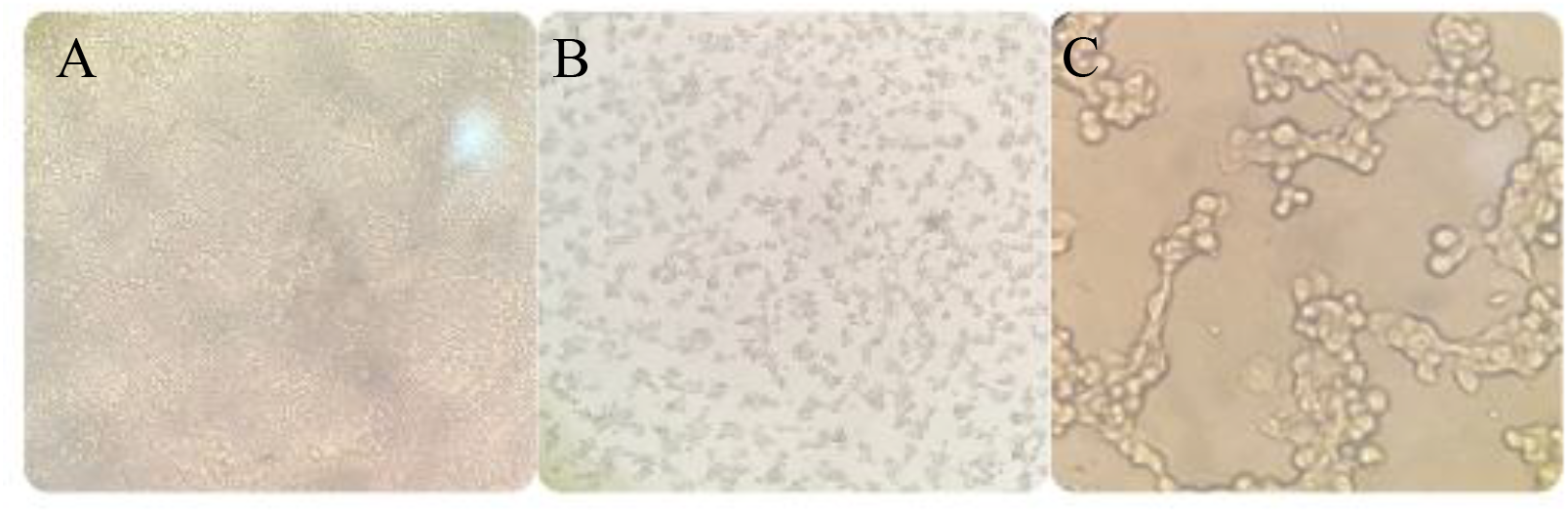
Light microscopy of cytopathic effects in Vero cells following 48 h of exposure to HBoV. (A) Uninfected Vero cells showing normal cellular morphology. (B) HBoV-infected Vero cells observed at 10× magnification. (C) Infected cells observed at 40× magnification. *Source: 2 Authors’ own work, 2025*.

**Figure 3.**
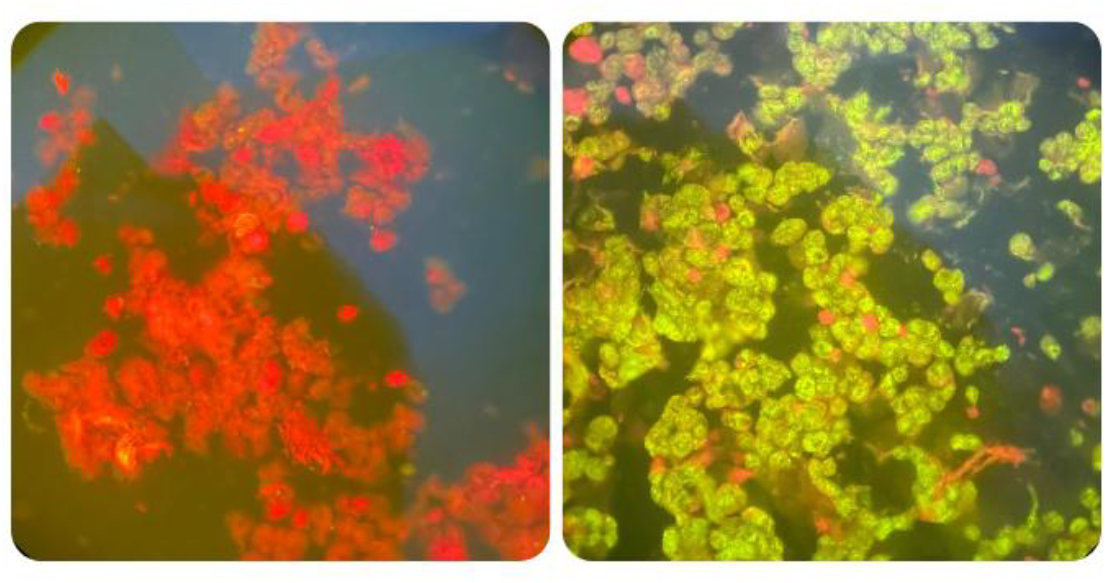
Vero cells visualized by fluorescence microscopy using a 40× objective. (A) Vero cells not infected with HBoV. (B) HBoV-infected cells. *Source: 3 Authors’ own work, 2025.*

Statistical analyses were conducted to evaluate the occurrence of HBoV infection in the study population and possible associations between viral detection and patients’ epidemiological characteristics. Variables included age, sex, age group, and presence of coinfections, with absolute and relative frequencies calculated. Age-related data are presented in Table 1. The reported mean age was 73 years among HBoV-positive patients and 56 years among HBoV-negative patients. The distribution of cases by sex is shown in Table 2.

Of the 178 gastroenteritis cases investigated, 10 (5.6%) tested positive for HBoV and 168 (94.4%) tested negative. Among female patients, 7 were HBoV-positive (6.36% of the 110 women), accounting for 70.0% of all positive cases, while 103 (93.64% of women) tested negative, representing 61.31% of all negative cases. Among male patients, 3 were HBoV-positive (4.41% of the 68 men), accounting for 30.0% of all positive cases, while 65 (95.59% of men) tested negative, representing 38.69% of all negative cases. Overall, 110 patients (61.8%) were female and 68 (38.2%) were male (Table 2).

**Table 1.** Frequency and percentage of AGE cases and HBoV-positive cases in the study population.

|  | AGE cases (n = 178) |  | HBoV-positive AGE cases (n = 10) |  |
| --- | --- | --- | --- | --- |
| Age group | Frequency | Percentage | Frequency | Percentage |
| **0-9** | 6 | 3.37% | 0 | 0% |
| **10-19** | 10 | 5.62% | 0 | 0% |
| **20-29** | 24 | 13.48% | 1 | 10% |
| **30-39** | 25 | 14.04% | 2 | 20% |
| **40-49** | 24 | 13.48% | 0 | 0% |
| **50-59** | 16 | 8.99% | 0 | 0% |
| **60-69** | 25 | 14.04% | 2 | 20% |
| **70-79** | 15 | 8,43% | 4 | 40% |
| **80-89** | 28 | 15.73% | 1 | 10% |
| **90+** | 5 | 2.81% | 0 | 0% |
| **Total** | 178 | 100% | 10 | 100% |

Source: 4 Authors' own work, 2025.

**Table 2.** Distribution of AGE cases by sex.

|  | Gastroenteritis cases |  |  |
| --- | --- | --- | --- |
| **SEX** | **HBoV-positive** | **HBoV-negative** | **Total** |
| **Female** | 7 | 103 | 110 |
| Within females | 6.36% | 93.64% | 100% |
| Share of all positive cases | 70.00% | N/A* | N/A* |
| **Male** | 3 | 65 | 68 |
| Within males (%) | 4.41% | 95.59% | 100% |
| Share of all positive cases | 30.00% | N/A* | N/A* |
| **TOTAL** | 10 | 168 | 178 |
| Overall percentage | 5.62% | 94.38% | 100% |

\* N/A: not applicable.
Source: 5 Authors' own work, 2025.

## Discussion

The AGE findings were similar to those previously reported in Brazil (Carolina et al., 2007), but differed from earlier results obtained in Salvador (Campos et al., 2016). The previously reported prevalence in Salvador was higher, whereas the prevalence observed in the present study was approximately 24-fold lower. This difference could indicate a decline in the incidence of HBoV among patients with AGE in the city. The findings were also similar to those reported in studies from other countries (Cheng et al., 2008; Lekana-Douki et al., 2018; Lin et al., 2020; Nora-Krukle et al., 2018).

The age distribution of HBoV-positive cases showed that most detections occurred in older adults, particularly those aged 70–79 years. This finding suggests that older individuals may be vulnerable to HBoV infection, potentially because of age-related factors, compromised immune function, or associated comorbidities.

The 30–39-year age group also accounted for a notable proportion of HBoV-positive cases. No positive cases were detected in the youngest age groups (0–9 and 10–19 years), which may suggest lower HBoV prevalence or impact among children and adolescents and could reflect differences in immune responses or viral exposure between age groups. Despite the low frequency observed among children, adults may transmit these infections to children, a population at risk. Children have immature immune systems and may develop dehydration rapidly; their lower body weight may also increase their vulnerability to infection (Brandt; de Castro Antunes; da Silva, 2015; Pijnacker et al., 2019).

During the study period, Salvador was experiencing a viral outbreak attributed to norovirus (NoV). The predominance of NoV in the diagnosis of AGE may mask HBoV infection and could help explain the low HBoV prevalence reported in some studies (Carolina et al., 2007; Cheng et al., 2008). However, coinfection with these viruses may worsen clinical outcomes (Kumar et al., 2016).

Previous studies have described HBoV as difficult to propagate in cell culture (Allander, 2008; Huang et al., 2012; Schildgen et al., 2008). Nevertheless, HBoV cultivation in the human colorectal adenocarcinoma cell line Caco-2 has been reported using samples from patients with gastroenteritis (Ghietto et al., 2017). The authors did not identify previous reports of HBoV cultivation in Vero cells and therefore describe the present findings as the first report involving this cell line. The cytopathic effects observed during isolation may result from cytoskeletal rearrangement and dysregulation of cellular signaling pathways, including pathways involved in apoptosis, during viral replication, ultimately leading to cell death (Knipe; Howley, 2013).

HBoV uses cellular receptors and host replication, transcription, and translation machinery to produce new infectious viral particles. HBoV replication is highly dependent on dividing cells, which may help explain the challenges of propagating this virus in vitro (Huang et al., 2019). Laboratory cell lines such as Vero cells may enter a quiescent (nondividing) state when maintained in culture for extended periods, potentially limiting their ability to support efficient HBoV replication.

Immunofluorescence analysis enabled visualization of viral antigens in the infected cells. The fluorescence pattern may be related to the HBoV replication cycle, because this DNA virus replicates in the host cell nucleus, where viral components are synthesized and processed. During infection, however, viral proteins may also be distributed among different cellular compartments, particularly during particle assembly and release. Accordingly, the observed fluorescence distribution should be interpreted in the context of different stages of viral replication rather than restricted to nuclear localization. As a complementary method, immunofluorescence therefore contributes to confirming the presence of the virus in cultured cells and to characterizing the infection process.

Western blot analysis demonstrated the presence of viral proteins in HBoV-infected cells. The observed bands had approximate molecular masses of 93, 68, and 60 kDa and may correspond to the capsid proteins VP1, VP2, and VP3, respectively. Published values for VP1, VP2, and VP3 are approximately 86 kDa, 64–75 kDa, and 60–62 kDa, respectively (Knipe; Howley, 2013). The discrepancies between the molecular masses observed in this study and those reported in the literature may reflect experimental conditions and differences in protein migration during electrophoresis. Immune responses to HBoV involve both B and T lymphocytes, with capsid proteins contributing to immune activation. VP1 may stimulate IgG antibody production, whereas VP2 has been associated with T-cell activation. Nevertheless, antibody production against HBoV has been particularly linked to VP2 (Bhat; Almajhdi, 2021).

## Conclusion

The findings indicate the continued detection of HBoV during AGE outbreaks in Salvador, with a reported incidence of 5.7%. This figure is consistent with some reports in the literature (Carolina et al., 2007) but lower than that reported in Salvador in 2012, suggesting a possible reduction in viral circulation. The successful isolation of HBoV in Vero cells represents a novel contribution, given the difficulties associated with cultivating this virus. This study highlights the importance of continued HBoV research and the development of more accurate diagnostic methods to reduce the health burden of HBoV-associated AGE and help limit viral transmission.

## Data Availability

All data produced in the present work are contained in the manuscript

